# Developing and testing two arts-based knowledge translation tools for parents about pediatric acute gastroenteritis

**DOI:** 10.1101/2021.06.08.21258514

**Authors:** Shannon D. Scott, Anne Le, Lisa Hartling

**Author notes:** **Author Contributions** This study was conducted under the supervision of Dr. Shannon D. Scott (SDS), NPI for **translation Evidence in Child Health to enhance Outcomes** (ECHO) Research. SDS led and supervised all aspects of tool development and evaluation. Lauren Albrecht (Project Coordinator) coordinated the research process and drafted some of the earlier versions of aspects of the findings. Lisa Hartling, Terry Klassen, and Lisa Knisley assisted with organization of data collection in rural and remote regions and assisted with script iterations. This work was funded by: **Canadian Institutes of Health Research** Scott, S.D., Knisley, L., Klassen, T., Hartling, L., Archibald, M., & Hamm, M. (2014). Knowledge translation tools for parents with children with croup and gastroenteritis. Canadian Institutes for Health Research (CIHR) Operating Grant: Knowledge to Action ($191,351). 2014-2016.

## Abstract

Characterized by vomiting, fever, abdominal pain and diarrhea, acute gastroenteritis (AGE) is a common illness in pediatric populations. In Canada, pediatric AGE accounts for 200,000 emergency visits, 20,000 hospital admissions, and 30 deaths a year. Yet, there continues to be significant practice variations in the treatment of AGE. Knowledge translation (KT) can help close the research-practice gap. In particular, art and stories are powerful mediums that cut across age, culture, language, literacy, and gender barriers.

The purpose of this study was to work with parents to develop an e-Book and whiteboard animation video for parents on pediatric AGE. Using a multi-method research process, we developed a 2 minute 57 second video and 39-page e-Book for pediatric AGE. Both tools underwent usability testing with parents in three Canadian emergency department waiting rooms in urban, rural, and remote regions. Focus groups were also conducted with parents in each of the three regions.

Overall, parents felt that digital and paper-based KT tools would be beneficial knowledge dissemination mediums. Our study showed that parents positively rated an e-Book and whiteboard animation video for pediatric AGE. These findings demonstrate how working together with key stakeholders can facilitate the development of KT tools for parents that are usable, relevant, and increase parental confidence. Furthermore, the type of KT tool developed is an important decision that may depend on parental preferences as well as when and where parents access the tools.

**This report should be cited as:** Scott, S.D., Le, A., Hartling, L. (2021). Developing and testing two arts-based knowledge translation tools for parents about pediatric acute gastroenteritis. Internal Technical Report. ECHO Research, University of Alberta.

*Available at: http://www.echokt.ca/research/technical-reports/*

## Introduction

Characterized by vomiting, fever, abdominal pain and diarrhea, acute gastroenteritis (AGE) is an important, potentially preventable cause of morbidity and mortality in young children [1-3]. It is estimated that AGE in Canadian children less than five years of age annually accounts for 200,000 emergency visits, 20,000 hospital admissions and approximately 30 deaths a year [4]. In addition to the obvious impact that AGE has on children, families and health care systems around the globe, there is considerable practice variation thereby further compounding the impact of this common childhood condition [2, 5]. In particular, much effort has been given to the implementation of inexpensive, easy-to-use oral rehydration solution for the treatment of dehydration, yet in spite of the large body of evidence supporting its safety and efficacy it continues to be underutilized [2].

Knowledge translation (KT) refers to the science of developing and evaluating strategies to close the research-practice gap. KT in child health is unique given the focus on family-centred care and the extent and level of parental involvement in all aspects of care. This demands that energies be focused on developing KT interventions or tools specifically for parents to communicate complex child health information and facilitate health decision-making.

The telling of stories is one of the oldest forms of communication across cultures [6]. Art is a highly accessible form that does not require specialized knowledge and skills to derive meaning. Together, stories and art are powerful mediums that can cut across age, culture, language, literacy and gender barriers. Independently and together, stories and art have the potential to impart knowledge, create understanding, and sustain interest by generating meaningful connections. The literature on the benefits of story and art as mediums for KT is growing. Our previous research in this area demonstrates the potential of arts and narrative based forms for communicating with and influencing individuals [7, 8]. These approaches are ideal to put research-based child health knowledge into action for parents, yet further implementation and evaluation is urgently needed to guide their application.

Both e-Books and whiteboard animation videos utilize story and art in unique ways. Typically, with e-Books, the story is the central focus with art and/or sound used to complement the narrative. With whiteboard animation videos, the central focus is graphic, dynamic drawings complemented by a voiced narrative to convey knowledge. Previous e-Book evaluation in child health has shown parents prefer creative means of receiving information over traditional methods, such as handouts or brochures [9-12]. Additionally, current evaluation of a whiteboard animation video for parents about childhood croup demonstrates good usability by parents in urban, rural, and remote settings and excellent uptake when available for free online. The purpose of this study was to develop, refine, and evaluate the usability of one e-Book and one whiteboard animation video for parents about pediatric AGE and employing narrative and arts elements.

## Methods

Through a multi-method research process, the following research question guided this study: What is the usability of an e-Book and a whiteboard animation video for parents on pediatric AGE? Research ethics approval was obtained from the University of Alberta Health Research Ethics Board [Pro00050107]. Additionally, ethics and operational approvals were obtained by individual hospitals to conduct usability testing.

### Compilation of Parents’ Narratives

Parental narratives were informed through semi-structured qualitative interviews (**Appendix A**) and a systematic review. Caregivers of a child with AGE were recruited for this study in the pediatric emergency department (ED) at the Stollery Children’s Hospital in Edmonton, Alberta. Individual interviews were conducted (n=15), and thematic analysis of interview transcripts was completed using a hybrid inductive/deductive approach. Results from the qualitative interviews are published elsewhere [13].

### Iterative Tool Development Process

Interview data informed the development of a composite narrative for both tools. Artists (i.e., writer, illustrator, animator, voice actor, etc.) were recruited through a competitive process. The final video was 2 minutes 57 seconds in length and highlighted the story of a young child affected by AGE whose symptoms were manageable at home. The video was narrated by the child’s mother and highlighted the families’ past experiences with AGE, which informed their decisions in the present (**Appendix B)**. The 39-paged e-Book, titled “Sick All Night: When to go to the Hospital with Childhood Vomiting and Diarrhea”, was written from the perspective of a mother whose twins develop different severities of AGE (**Appendix C**). Both tools provided parents with details on what to expect when a child has AGE, research informed management strategies, and instructions on what to do if a child’s condition does not improve. Bottom Line Recommendations (BLR) for AGE developed by TRanslating Emergency Knowledge for Kids (TREKK) were integrated into the tools [14].

### Quantitative Usability Data

Parents were recruited to participate in an electronic, usability survey in three Canadian ED waiting rooms representing urban (Stollery Children’s Hospital), rural (Portage District General Hospital), and remote (Stanton Hospital) health regions. Members of the study team approached parents in the ED to determine interest and study eligibility. Study team members were available in the waiting rooms to provide technical assistance and answer questions as parents were completing the surveys. Surveys were informed by a systematic review of over 180 usability evaluations and comprised of 8, 5-point Likert items assessing: 1) usefulness, 2) simplicity, 3) levels of engagement, 4) satisfaction, 5) quality of information, and 6) perceived value (**Appendix D**). Data were analyzed using SPSS v. 24 [15]. Parents were also asked to provide their positive and negative opinions of the tool via two free text boxes. Focus groups were audio-recorded and transcribed verbatim. Transcripts were cleaned and analyzed using NVivo 11 [16].

### Qualitative Usability Data

Parents were purposively recruited to participate in one semi-structured focus group in each of the three regions. In two regions, parents were recruited from existing parent advisory groups involved in child health research. In the third region, the local public health unit distributed flyers about the focus group to their parent groups. The interview consisted of questions about: a) tool viewing experience, b) tool attributes that were useful, c) tool elements that were not helpful, d) perceptions of the tool’s utility, and e) recommended revisions (**Appendix E**). Key aspects of the tools, including narrative, visual appear, health information, length, engagement and interactivity, were explored. Focus groups were audio recorded on secured iPads and digital recordings were transcribed verbatim. Data was analyzed deductively using the semi-structured interview guide to develop broad categories. Focus group data were then coded, codes were placed into the broad categories, and the categories synthesized into themes.

The timeline for this project can be viewed in **Appendix F**.

## Results

### Usability Survey Results

Sixty-three surveys were completed by parents in the three sites. Participant demographics are presented in Table 2. Usability measures were assessed with a 5-point Likert scale with 1 being the lowest (i.e., most negative) option and 5 being the highest (i.e., most positive) response. The overall results of the usability survey measures are presented in Table 3. Table 4 presents usability survey results by region (urban vs rural/remote).

**Table 1.**
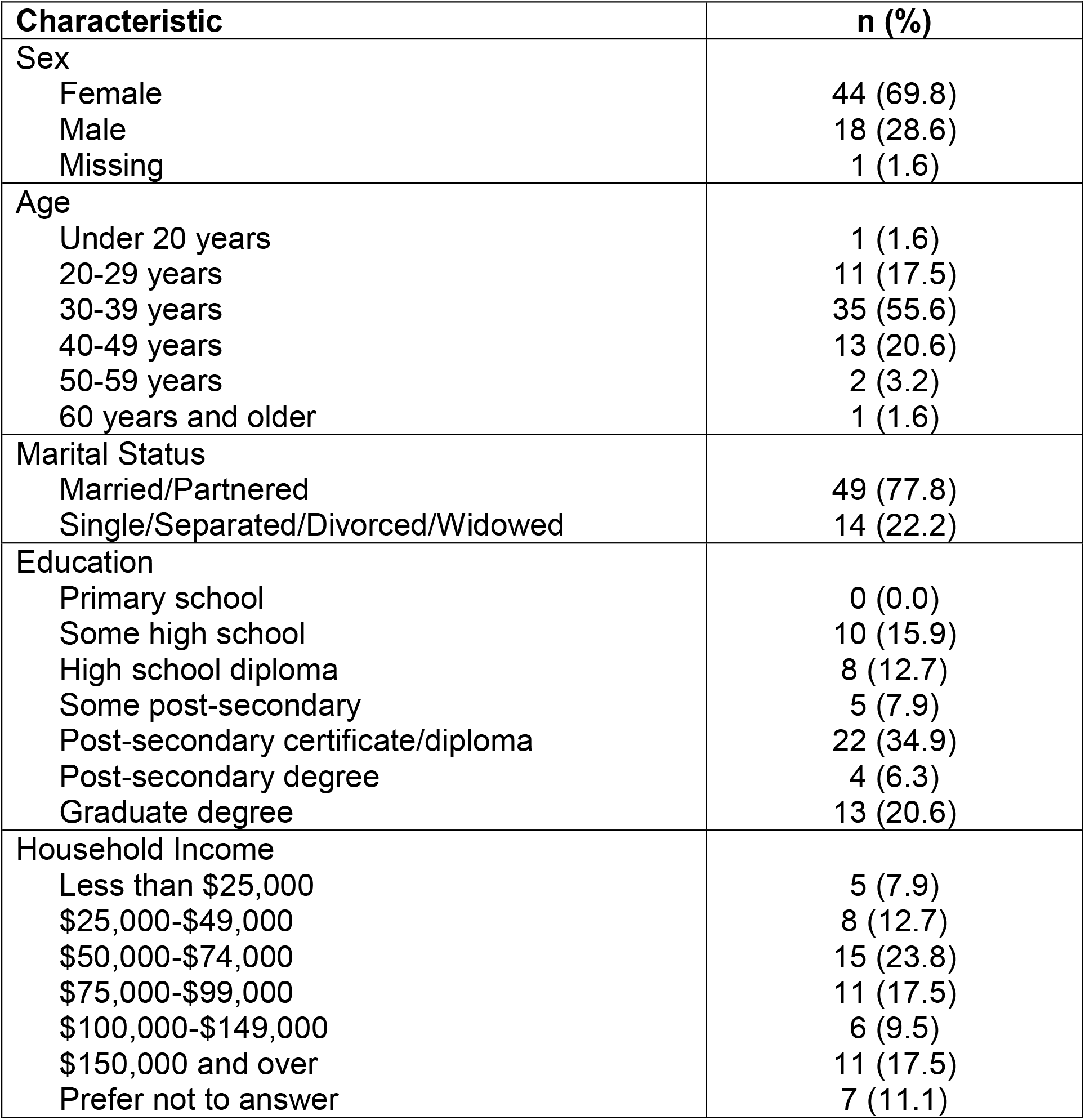
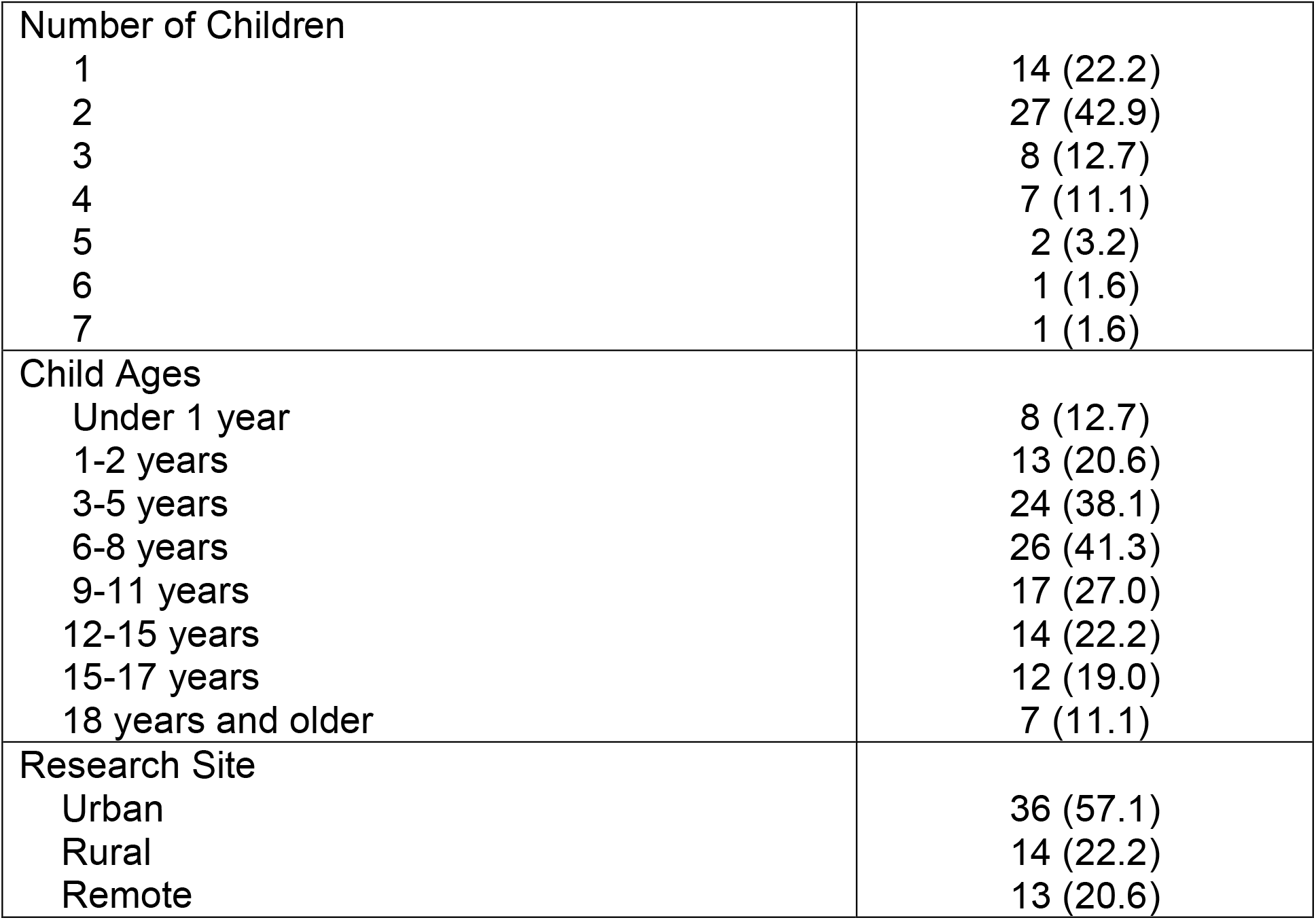
Demographic characteristics of parents who assessed the usability of the whiteboard animation video (N=63)

**Table 2.** Means of participant responses to the usability survey.

| Usability Measures | Video | E-Book |
| --- | --- | --- |
| It is useful. | 4.38 (0.779) | 4.29 (1.013) |
| It meets my information needs. | 4.38 (0.817) | 4.04 (1.105) |
| It is simple to use. | 4.56 (0.960) | 4.03 (1.349) |
| I can use it without written instructions or additional help. | 4.35 (1.152) | 4.17 (1.311) |
| It is fun to use. | 4.26 (0.898) | 3.38 (1.321) |
| I am satisfied with it. | 4.62 (0.604) | 4.10 (0.604) |
| I would use it in the future. | 4.41 (1.019) | 3.45 (1.429) |
| I would recommend it to a friend. | 4.67 (0.736) | 4.21 (1.114) |

**Table 3.** Demographic characteristics of parents who participated in focus group discussions (N=19)

| **Characteristic** | **n (%)** |
| --- | --- |
| Sex |  |
| Female | 11 (57.9) |
| Male | 8 (42.1) |
| Age |  |
| 20-29 years | 3 (15.8) |
| 30-39 years | 4 (21.1) |
| 40-49 years | 10 (52.6) |
| 50-59 years | 1 (5.3) |
| 60 years and older | 1 (5.3) |
| Marital Status |  |
| Married/Partnered | 13 (68.4) |
| Single/Separated/Divorced/Widowed | 5 (31.6) |
| Education |  |
| Primary school | 0 (0.0) |
| Some high school | 0 (0.0) |
| High school diploma | 1 (5.3) |
| Some post-secondary | 1 (5.3) |
| Post-secondary certificate/diploma | 3 (15.8) |
| Post-secondary degree | 10 (52.6) |
| Graduate degree | 2 (21.1) |
| Household Income |  |
| Less than \$25,000 | 3 (15.8) |
| \$25,000-\$49,000 | 2 (10.5) |
| \$50,000-\$74,000 | 2 (10.5) |
| \$75,000-\$99,000 | 2 (10.5) |
| \$100,000-\$149,000 | 3 (15.8) |
| \$150,000 and over | 5 (26.3) |
| Prefer not to answer | 2 (10.5) |
| Number of Children |  |
| 1 | 4 (21.1) |
| 2 | 7 (36.8) |
| 3 | 3 (15.8) |
| 4 | 4 (21.1) |
| Research Site |  |
| Urban | 8 (42.1) |
| Rural | 4 (21.1) |
| Remote | 7 (36.8) |

### Focus Group Findings

Nineteen parents participated in 3 focus groups; one focus group in each of the three regions. The average number of participants was six per focus group. Participant demographics are presented in Table 3. Focus group length ranged from 23 minutes to 42 minutes.

The e-Book was described by parents as looking attractive (i.e., colouring, illustrations) and containing relevant, helpful information. However it was deemed by most participants to be too long with dense text on the pages. Some found the narrative elements distracting, *“She kept going back to her cleaning. I’m like ‘okay’. This is it. Sorry. But I wouldn’t be doing that.” (Participant 1, focus group 2)*, but others felt that the human aspects of the story resonated with them and provided comfort *“But if you’re in the Emergency Room, and you’re waiting and waiting – it’d be nice to read that for a while” (Participant 1, focus group 3)*. Another major issue identified was that the target audience for the e-Book was unclear. Participants indicated that the storybook appearance (i.e., colours, illustrations) were more suitable for a child audience, but the story content was too complex for a child and would be better suited for parents. *“I think the art work here is beautiful […] it’s just a really, really nice potential, to make it into a story for a kid” (Participant 6, focus group 2)*. Ultimately participants agreed that the e-Book was too long to be read when the child was sick and the parents were trying to determine the best course of action and that it would be better provided as anticipatory guidance or as informative material provided in clinics or emergency department waiting rooms. *“At page 13, I was like, nah… this is more than I can do, especially if I’m in a moment where I need the information right away” (Participant 1, focus group 1). “I found I kind of lost interest part of the way through and, so I don’t think it got me the information as quickly as I wanted it to, I think” (Participant 3, focus group 1)*.

The whiteboard video was described as informative, clear, and to-the-point. Participants indicated that the video reflected real life and that the checklists would be very helpful when making a decision about how best to care for a sick child. Participants liked the video visuals, but some felt more colour would increase appeal and others were distracted by the hand conducting the drawing. *“I liked everything about it. I love that she read from a check list. I just really liked it… I just really like the fact that the mom was reading from you know, this checklist – like she’s explained like you know, she had a doctor’s plan but then the checklist was there, ‘cause she – you know, this was a previous thing as baby – I really liked that. It just really felt real.” (Participant 3, focus group 1)*.

Participants agreed that digital and paper-based KT tools have their place. As one participant described, *“But if I was panicking, I want the video. If I had time, like waiting room or… just browsing, I like the e-book better. ‘Cause then I can get kids involved.” (Participant 4, focus group 3)*. Parents also stressed that the context in which they watch the video is important. *“I think it makes a difference what the context is. Like, when I’m sitting, waiting in the doctor’s office, ah, if I see a video that sort of starts with the back story, I’ll sit back. I’ll say “oh this is interesting. If I watch it, maybe I’ll get educated’. But if you’re a parent who’s in the middle of a crisis situation, you’re probably gonna get to the point” (Participant 7, focus group 2)*. When asked whether they had any clear preferences for e-Book or video, participants agreed that preferences were situational, and one parent answered: *“I guess it depends where you expect people to see it. Are they supposed to watch this in the Emergency room, in their doctor’s office? ‘Cause… depending on where you want them to be viewing this, it kinda… changes how you… how you assess it” (Participant 4, focus group 2)*. However, many participants also indicated that hard copy materials (i.e., pamphlets) are easily misplaced or recycled. For the e-Book there were many participants who agreed that this type of story might be better presented as a hard copy, children’s story. According to another participant, *“Um, but I like the idea of a hard copy of a story book that I can read to the child while I’m there [in the Emergency Department]. You know, so it’s less scary. ‘Cause it’s one thing for me to say, ‘hey, this isn’t scary.’ But to sit there and read, you know, a story book that is reassuring and comforting, I wouldn’t mind that.” (Participant 2, focus group 1)*.

### Conclusions

This study utilized a novel approach in the development and evaluation of an AGE knowledge translation tool for parents. We employed qualitative and quantitative methods to develop and evaluate 2 different KT tools for parents with a child with gastroenteritis. We engaged with stakeholders throughout the entire process by conducting interviews with parents prior to tool development, evaluating our tool through consultations with health care providers and parents, and qualitatively assessing our tool through parent interviews and open text survey questions.

Our findings demonstrate how working together with key stakeholders can facilitate the development of KT tools for parents that are usable, relevant, and increase parental confidence. Study findings also highlight the type of KT tool developed is an important decision that may depend on parental preferences as well as when parents access tools. Going forward, future research needs to explore when in the care trajectory and what types of situations demand particular types of KT tools.

Overall, the assessment of our tools was quite positive from parents and health care providers and provided constructive suggestions on how to improve the tools to better meet the information needs of parents. These findings highlight the positive impact of engaging end-users throughout knowledge translation research processes and suggest that art and narrative-based tools are highly usable and accepted by parents caring for children with acute illness.

**The tools can be found here: http://www.echokt.ca/tools/stomach-flu/**.

*Please note that as of the publication of this technical report, the video is not available as it is being evaluated in a feasibility trial*.

Note: Our KT tools are assessed for alignment with current, best-available evidence every two years. If recommendations have changed, appropriate modifications are made to our tools to ensure that they are up-to-date.

## Data Availability

All data from this project are stored securely at the University of Alberta and are only to be accessed by the study team to ensure participant privacy and confidentiality.

## Other Outputs from this Project

## Appendices

### Appendix A Qualitative Interview Guide

1. Tell me about your child that was ill. How old is your child? How was your child ill? Has your child previously had gastroenteritis (vomiting & diarrhea)?
2. Tell me about your experience having your child experience gastroenteritis/vomiting & diarrhea.
3. How did you feel during this experience?
4. What did you do to manage diarrhea and vomiting with your child? (any techniques that you used, for example, giving oral fluids, talking with family/friends etc.)
5. What strategies were put in place by health care professionals to help your child? (for example, did they use oral rehydration? Did they use IV fluids?) Did they ask you to do anything?
6. How did your child manage the experience? How did you feel about the outcome of this situation?
7. What did you learn from this experience?
8. If presented with the same situation again, would you do anything differently?

### Appendix B Video

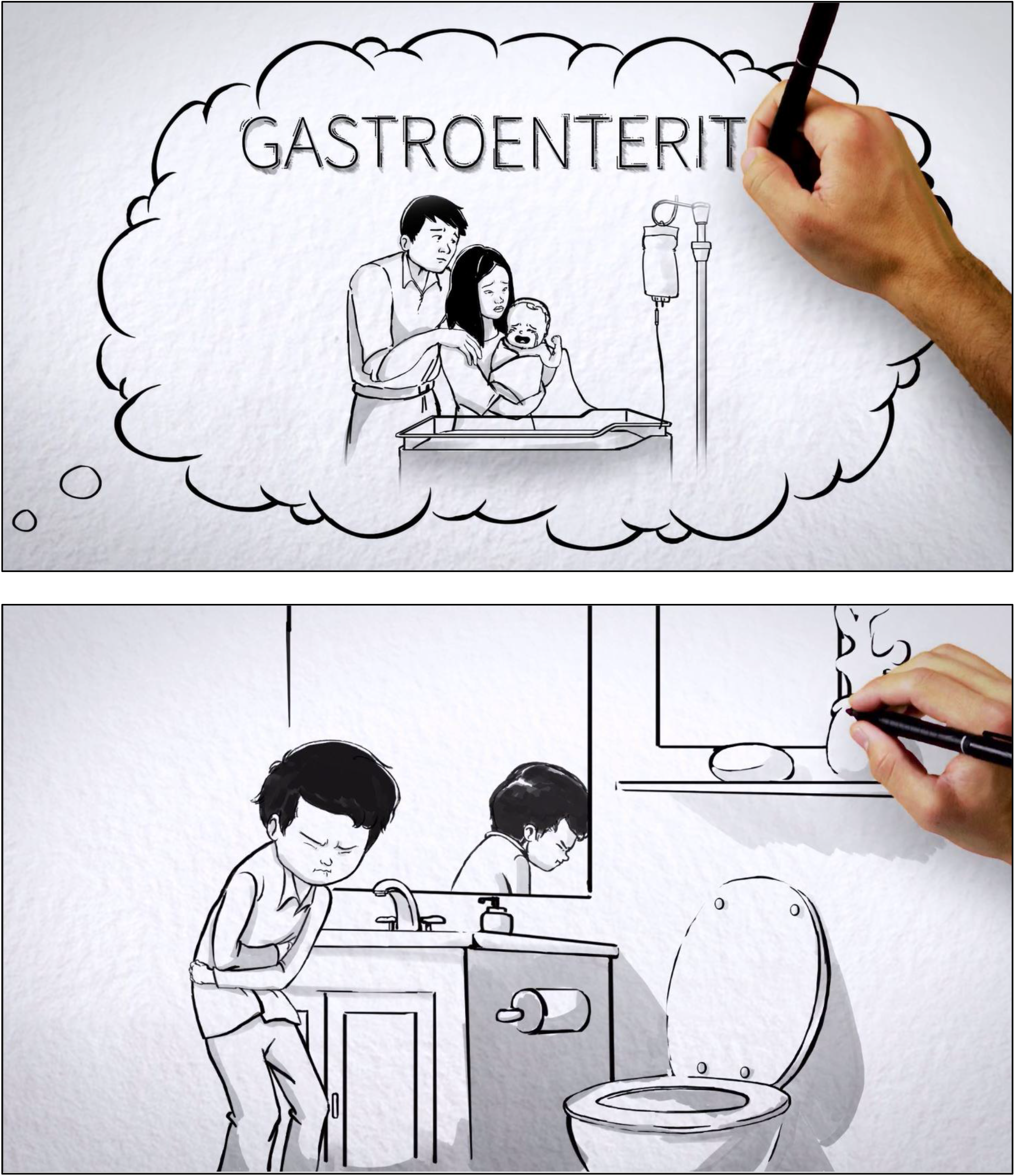
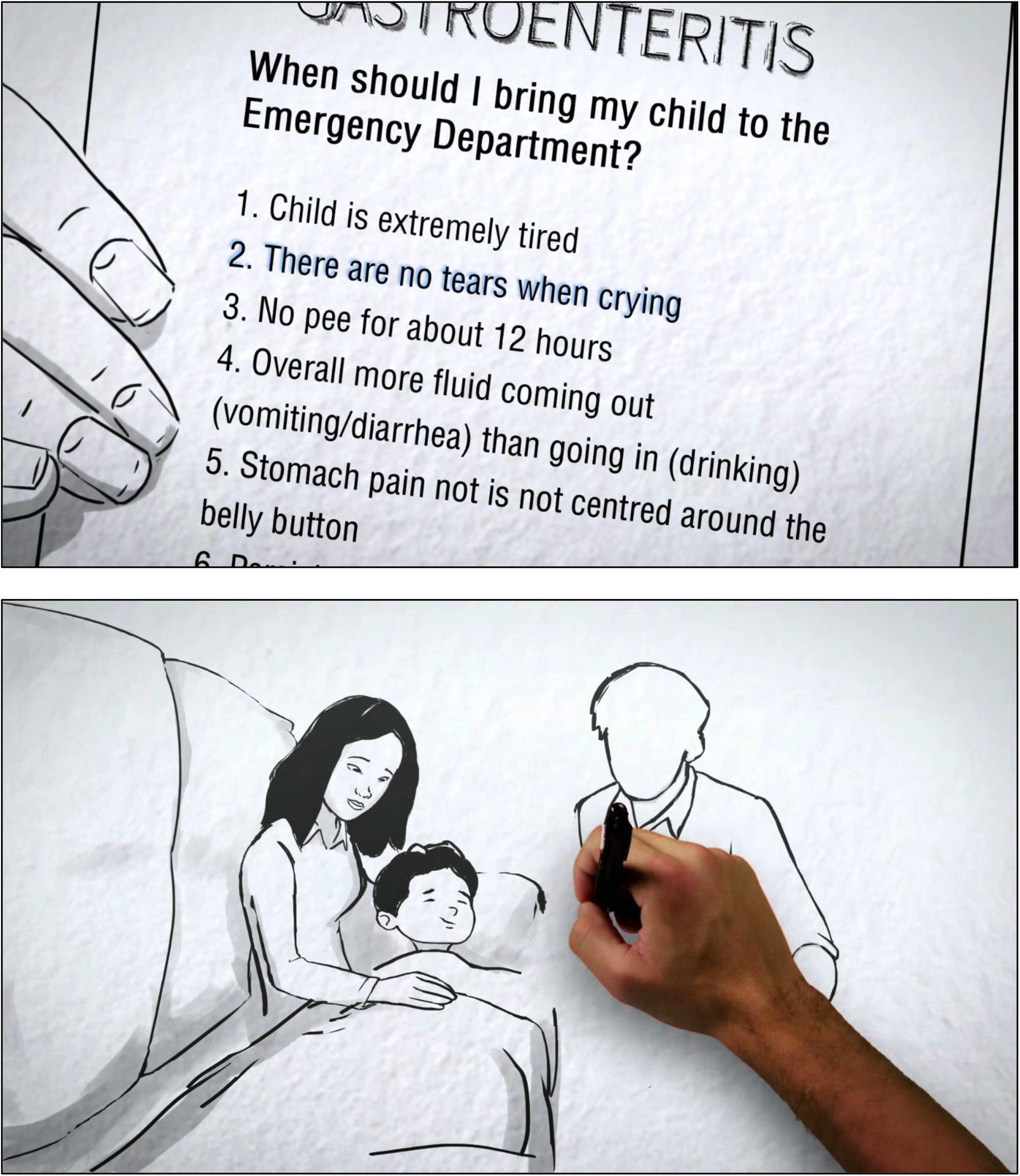

### Appendix C Gastroenteritis eBook

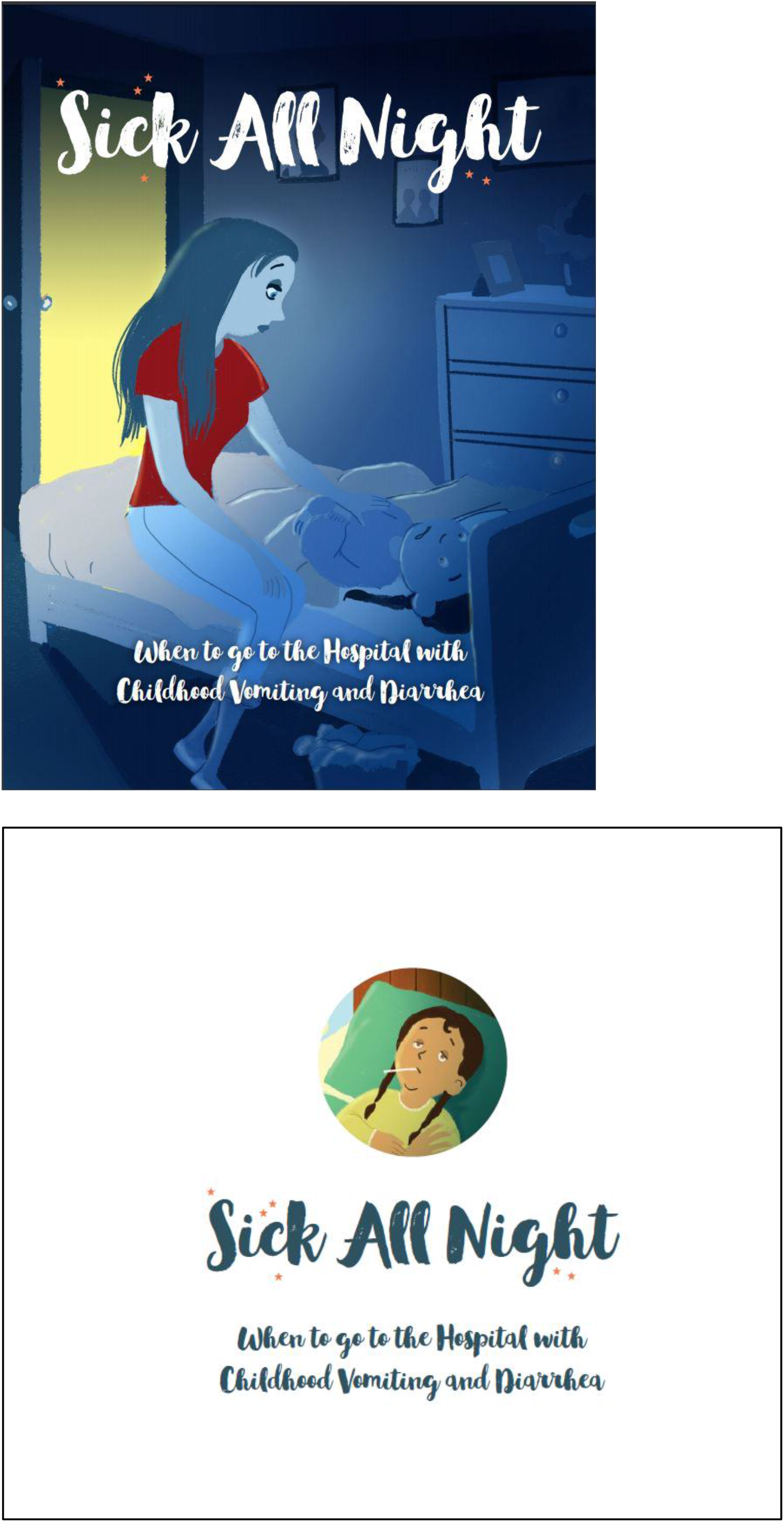
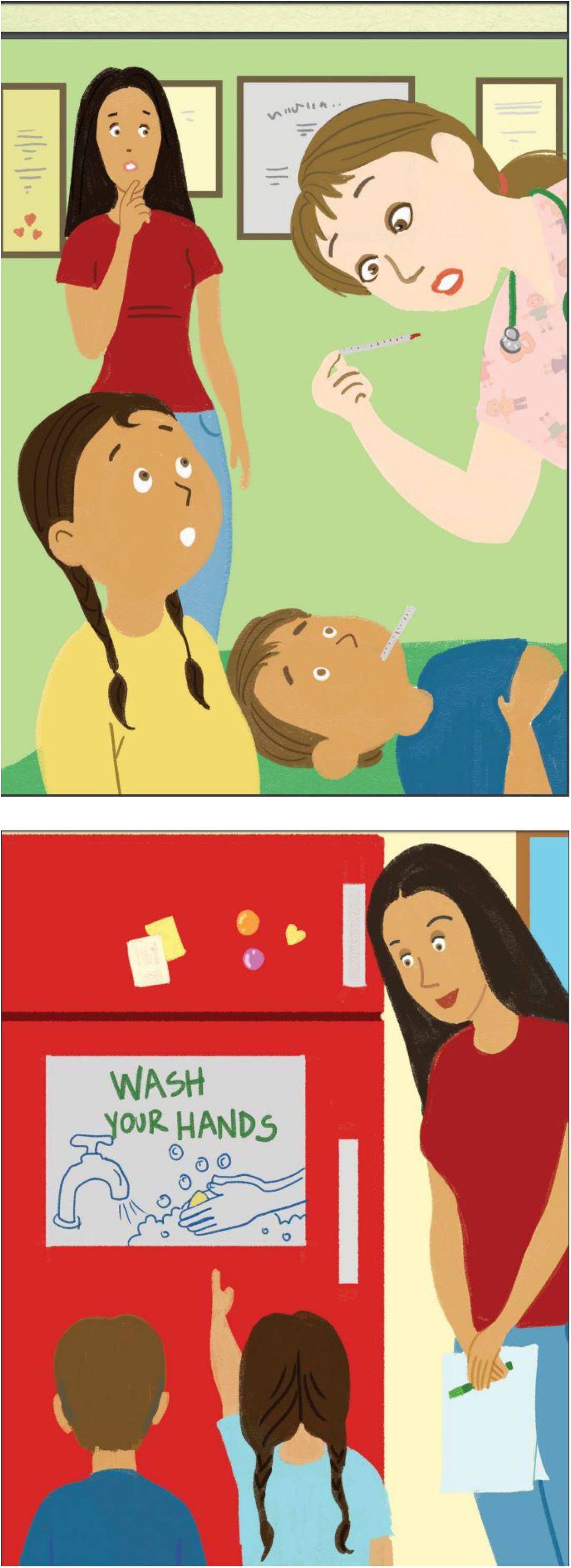

### Appendix D Usability Testing Survey

SECTION 1: Demographics

1) What is your gender?
  □ Male
  □ Female
3) What is your Age?
  □ Less than 20 years old
  □ 20-30 years
  □ 31-40 years
  □ 41-50 years
  □ 51 years and older
4) What is your Marital Status?
  □ Married
  □ Single
5) What is your gross annual household income?
  □ Less than $25,000
  □ $25,000-$49,999
  □ $50,000-$74,999
  □ $75,000-$99,999
  □ $100,000-$149,999
  □ $150,000 and over
6) What is your highest level of education?
  □ Some high school
  □ High school diploma
  □ Some post-secondary
  □ Post-secondary certificate/diploma
  □ Post-secondary degree
  □ Graduate degree
  □ Other
7) How many children do you have?_________
8) How old are your children?_________

SECTION 2: Assessment of attributes of the arts-based, digital tools

***\*participant is randomized to view 1 of 2 digital tools then automatically directed to the survey***

1. It is useful. [5-point Likert Scale]
2. It meets my information needs. [5-point Likert Scale]
3. It is simple to use. [5-point Likert Scale]
4. I can use it without written instructions or additional help. [5-point Likert Scale]
5. It is fun to use. [5-point Likert Scale]
6. I am satisfied with it. [5-point Likert Scale]
7. I would use it in the future. [5-point Likert Scale]
8. I would recommend it to a friend. [5-point Likert Scale]
9. List the most negative aspects: [open text]
10. List the most positive aspects: [open text]

### Appendix E Focus Group Guide

Good morning/afternoon. Thank you for taking the time to meet with us. We would like to ask you several questions about your impressions of the eBooks and Whiteboards. Our conversation is being transcribed to ensure that we have an accurate summary of your opinions. All the information we collect will be kept confidential. You may refuse to answer any questions or leave the focus group at any time. Do you have any questions before we begin? Please feel free to ask questions at any time during the interview.

Let’s get started;

1. Tell us your <u>first impressions</u> of the arts-based digital tools provided to you in advance of this focus group.
  - eBooks
  - Whiteboards
  - Describe the <u>usability</u> of these digital tools
2. Did the tools provide you with useful information about pediatric croup OR pediatric gastroenteritis?
  - Explain how
  - Tell me what information you learned.
  - If yes, what information was most useful? Least useful?
  - If no, what information was missing or inadequate?
3. What <u>new</u> information did you learn?
  - Difference between eBooks and Whiteboards
4. How do you anticipate that this information will influence your experience in the future?
5. How will the information provided in the digital tools help you make decisions?
  - Difference between eBooks and Whiteboards
6. How did accessing information in this format (eBooks and Whiteboards) compare with the more traditional parent educational sheets? What digital tool did you prefer?
7. Overall, what is your impression of the eBook? Why did you like it? Why did you dislike? <u>prompt</u>
  ▪ Story/narrative
  ▪ Art
  ▪ Aesthetics/Style
  ▪ Size
  ▪ Color
  ▪ Format
  ▪ Readability
  ▪ Interactivity/Engagement
  ▪ Embedded audio
  ▪ Length
8. Overall, what is your impression of the whiteboard? Why did you like it? Why did you dislike? <u>prompt</u>
  ▪ Drawing style
  ▪ Story/Narrative
  ▪ Size
  ▪ Color
  ▪ Format
  ▪ Voice of the narrator
  ▪ Length
  ▪ Interactivity/Engagement
9. Are there any recommendations for additions, changes to the digital tools?
10. Would you recommend this digital knowledge translation tools to other parents?

Thank you for your thoughtful responses to our questions. Are there any other comments/concerns about the digital tools that have not yet touched upon?

### Appendix F Project Timeline

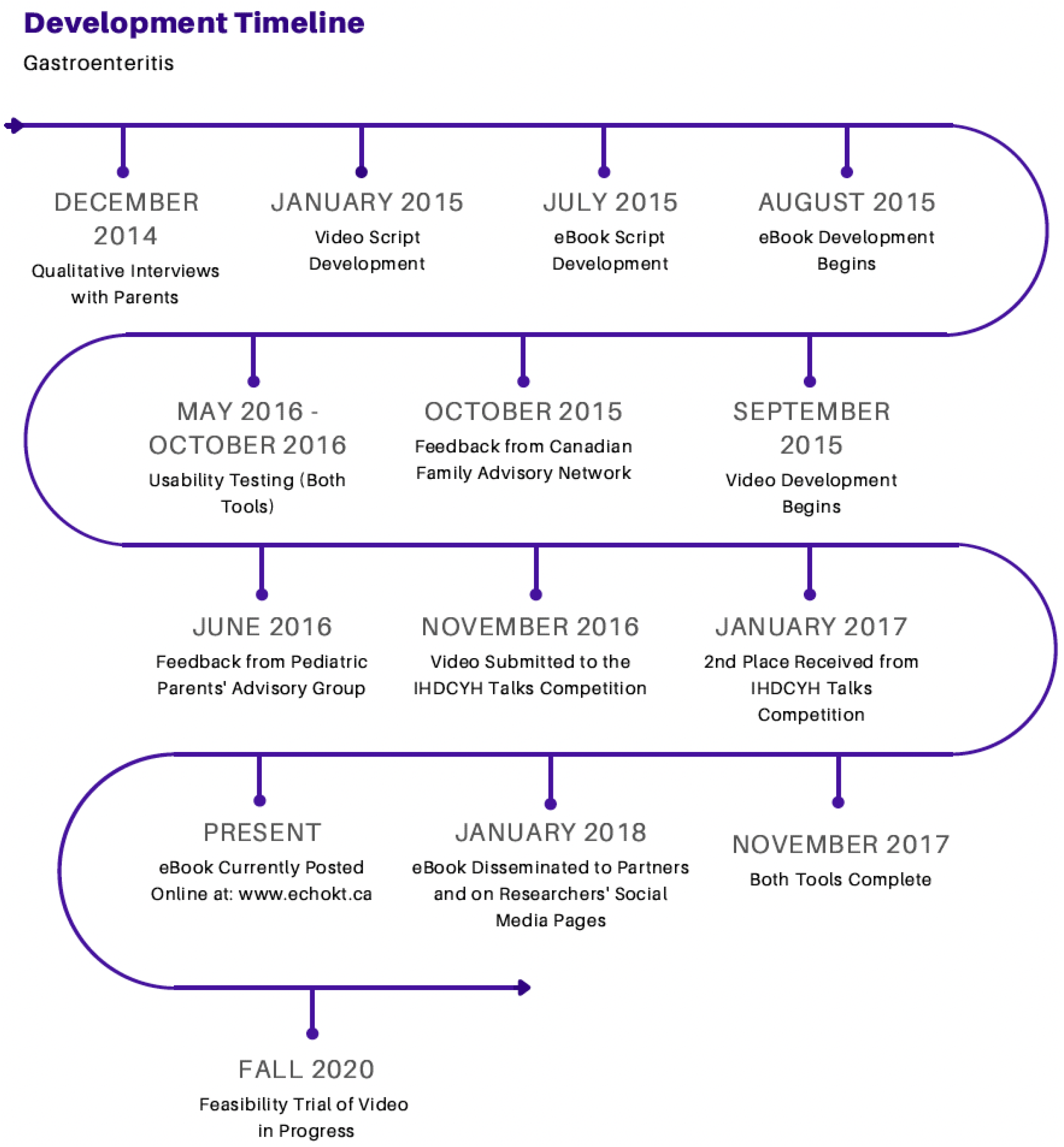

